# An acute bleomycin inflammatory and fibrotic mouse model of morphea is dependent upon CXCL9 and CXCR3

**DOI:** 10.1101/19000844

**Authors:** Jillian M. Richmond, Dhrumil Patel, Tomoya Watanabe, Colton J. Garelli, Madhuri Garg, Karen Dresser, April Deng, Carol A. Feghali-Bostwick, John E. Harris, Heidi Jacobe

## Abstract

Morphea, or localized scleroderma, is characterized by an inflammatory phase followed by cutaneous fibrosis, which may lead to disfigurement and/or disability. Previous work from our group showed that the CXCR3 ligands CXCL9 and CXCL10 are highly upregulated in lesional skin of morphea patients. Here, we used an acute inflammatory and fibrotic bleomycin mouse model of morphea to examine the role of the CXCR3 chemokine axis in pathogenesis. We first characterized which cells produce the CXCR3 ligands in the skin using the <u>R</u>eporter of <u>E</u>xpression of C<u>X</u>CR<u>3</u> ligands mouse (REX3). We found that fibroblasts contribute the bulk of CXCL9 and CXCL10, whereas endothelial cells are key dual chemokine producers. Macrophages, which have high MFI of chemokine expression, upregulated CXCL9 production over time, fibroblasts CXCL10 production, and T cells dual chemokine expression. To determine whether bleomycin treatment could directly induce expression of these chemokines, we treated cultured REX3 mouse dermis monolayers *in vitro* with bleomycin or IFNγ with TNF and found that bleomycin could induce low amounts of CXCL9 directly in fibroblasts, whereas the cytokines were required for optimal CXCL9 and CXCL10 production. To determine whether these chemokines are mechanistically involved in pathogenesis, we induced fibrosis in CXCL9, CXCL10, or CXCR3 deficient mice and found that fibrosis is dependent on CXCL9 and CXCR3. Addition of recombinant CXCL9, but not CXCL10, to cultured mouse fibroblasts induces collagen 1a1 mRNA expression, indicating the chemokine itself can contribute to fibrosis. Taken together, our studies provide evidence that acute intradermal bleomycin administration in mice can model inflammatory morphea, and that CXCL9 and its receptor CXCR3 are mechanistically involved in pathogenesis.

**One Sentence Summary:** CXCL9 drives acute morphea pathogenesis in mice.

## Introduction

Morphea, or localized scleroderma, is an autoimmune disease affecting the dermis and subcutis that results in fibrosis. In severe cases, fibrotic morphea can cause joint contractures, limb length discrepancy, and facial hemi-atrophy leading to disfigurement and disability (Fett and Werth 2011a). Inflammation is thought to precede and likely drive cutaneous fibrosis based on clinical observations, but the molecular events underlying these observations are poorly studied. Further, a lack of validated animal models of morphea has further hindered studies examining the molecular underpinnings of morphea (Laxer and Zulian 2006)(Martini et al. 2018). As a result, even though targeted, less toxic treatments are in development for many autoimmune diseases, none have been developed for morphea: the mainstay of morphea therapy remains non-specific immunosuppressants, specifically methotrexate and corticosteroids(Fett and Werth 2011b), the use of which is limited by side effects and toxicity. Studies to better understand the immune dysregulation in morphea are needed to identify targets for therapeutic development.

Our previous work, as well as others, has shown that the interferon (IFN)-inducible chemokines CXCL9 and CXCL10 are highly expressed in lesional skin and serum from morphea patients, and may serve as biomarkers for disease activity and severity (Mertens et al. 2018; Mirizio et al. 2019; O’Brien et al. 2017). Together with CXCL11, CXCL9 and CXCL10 bind to a shared receptor, CXCR3 (Colvin et al. 2004). The CXCR3 chemokine family primarily drives T cell migration during inflammation (reviewed in (Groom and Luster 2011)). Interestingly, CXCR3 has also been shown to be expressed on fibroblasts and proposed to mediate wound healing (Kroeze et al. 2012). CXCR3-deficient mice exhibit delayed re-epithelialization (Yates et al. 2009), decreased scar formation (Yates et al. 2010), and dermal differentiation defects (Yates et al. 2007), indicating activation of this receptor on fibroblasts by CXCL9 and CXCL10 may play a role in extracellular matrix formation and collagen deposition.

Mouse models of fibrosis have been based on injection of bleomycin, a chemotherapeutic with a known side effect of inducing fibrosis. While intravenous bleomycin in mice induces scleroderma-like disease and intratracheal administration induces a condition similar to idiopathic pulmonary fibrosis (Lam et al. 2016), subcutaneous injection has been shown to induce skin fibrosis with inflammatory infiltrates at early time points (Castelino et al. 2011; Yamamoto et al. 1999). We used a modified protocol of acute subcutaneous injection of bleomycin for 5 or 12 days to model inflammatory and fibrotic aspects of morphea.

Based on our preliminary studies showing increased CXCL9 and CXCL10 in the lesional skin of morphea patients and published literature suggesting CXCR3 plays a role in wound healing responses, we hypothesized that this chemokine family may play a role in morphea pathogenesis. We set out to test this using the acute subcutaneous bleomycin mouse model. Here, we show which cell types in the skin produce CXCL9 and CXCL10 using a novel reporter of expression of CXCR3 ligands mouse (REX3, (Groom et al. 2012)). To address the role that these chemokines play in morphea pathogenesis, we tested chemokine deficient mice and found that CXCL9 and CXCR3-deficient animals, but not CXCL10-deficient animals, were protected from dermal thickening and collagen deposition compared to WT controls. Finally, we treated mouse fibroblasts *in vitro* with recombinant CXCL9, and found that CXCL9 can dose-dependently increase transcription of collagen 1a1. Together, these studies lay the foundation for translational research in morphea, and further elucidate potential pathogenesis directed treatment approaches targeting both inflammation and fibrosis.

## Results

### Acute subcutaneous bleomycin induced inflammation and fibrosis in mice resembles human morphea

To induce morphea-like disease in mice, we performed subcutaneous injection of 100 mg of bleomycin in 2 locations on depilated dorsal back skin of C57BL/6J mice, and compared these mice to vehicle injected controls at day 5 and day 12 (Fig 1A). Notably, areas that were injected with bleomycin remained depilated, whereas areas that were injected with PBS regrew hair (white arrows, Fig 1B). Inflammatory infiltrates increased in the skin as determined by flow cytometry analysis of total CD45+ cells (Fig 1C). We compared the mouse histology to human morphea skin biopsies and found thickening of collagen bundles in the dermis to be similar, as assessed using Trichrome staining and similar patterns of inflammation (Fig 1D & E). In contrast, we found no obvious evidence of lung fibrosis on histology (Fig S1). Therefore, we concluded that this model of acute bleomycin administration resembled skin-limited rather than systemic sclerosis.

**Fig. 1.**
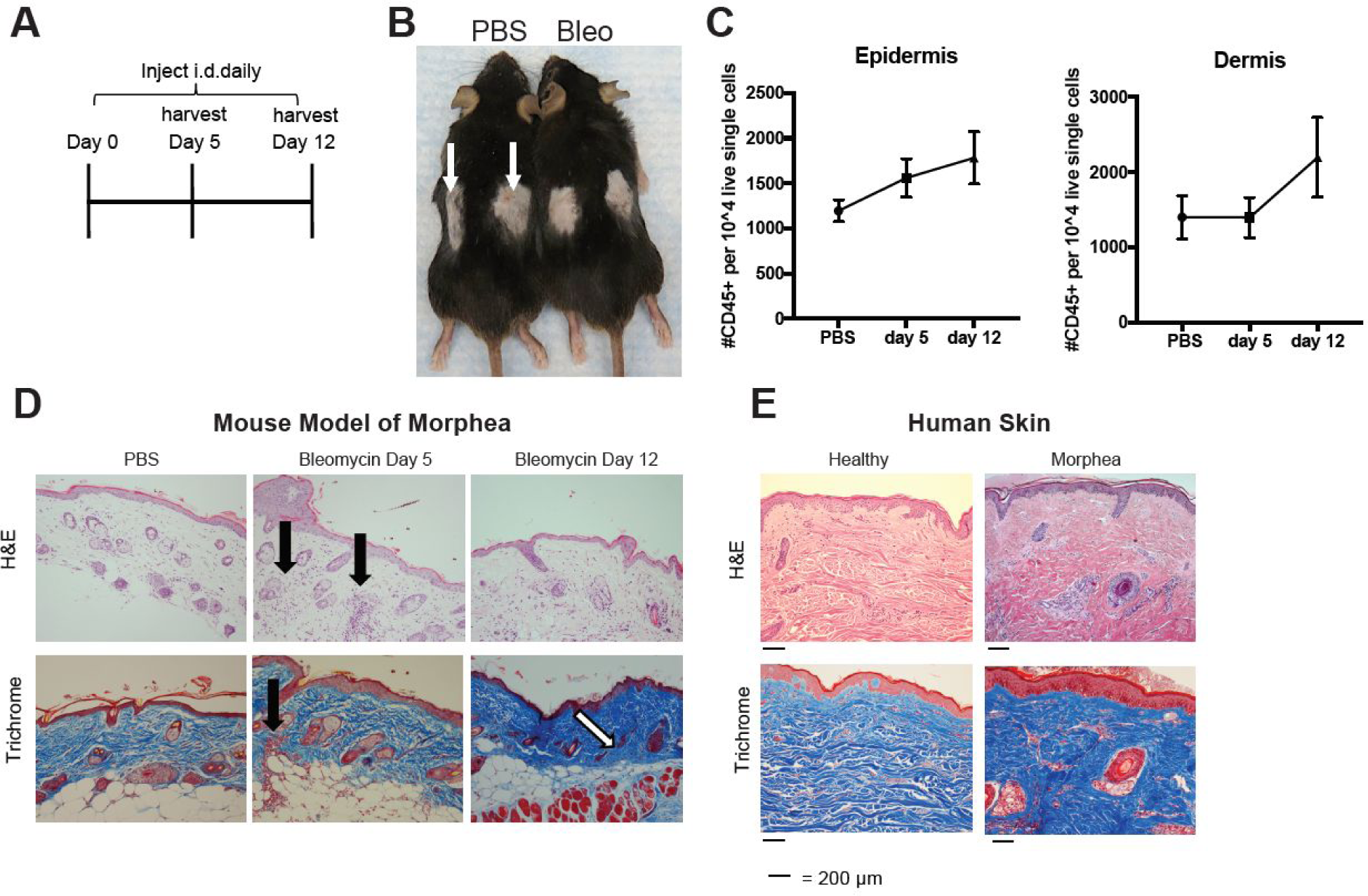
Acute administration of subcutaneous bleomycin induces fibrosis in mice with limited hair regrowth. (**A**) Model used to induce morphea in mice. (**B**) Sample photographs of injection sites on mice (white arrows indicate hair regrowth, which was seen less often in bleomycin treated areas compared to PBS treated areas). (**C**) Flow cytometry for live single CD45+ cells in the epidermis and dermis were normalized to 10,000 live singlets, and infiltrates were observed to increase by day 12. (**D**) Example H&E and Trichrome staining from the skin of mice treated with acute bleomycin. Black arrows indicate inflammatory infiltrates, white arrow indicates fat layer loss (100x light microscopy). (**D**) Example H&E and Trichrome staining from morphea patient skin (left panels 40x, right panels 100x light microscopy).

### T cells, antigen presenting cells and fibroblasts are increased in the skin in the fibrosis model

Next, we characterized the inflammatory infiltrates and stromal cells present in the skin of mice treated with bleomycin (see Table S1 for antibodies and flow panels). Epidermis and dermis were separated via enzymatic digestion, and single live cells were assessed for expression of cell surface markers (Fig 2A). We found significant increases in numbers of epidermal T cells (Fig 2B), dermal T cells, and dermal antigen presenting cells (APCs, including populations of CD11b+CD11c+ and CD11b+ macrophages and CD11c+ dendritic cells) but not B cells (Fig 2C). These findings mirror those found in human morphea lesions, namely increased T cell and macrophage signatures (Higashi-Kuwata et al. 2009; Mirizio et al. 2019).

**Fig. 2.**
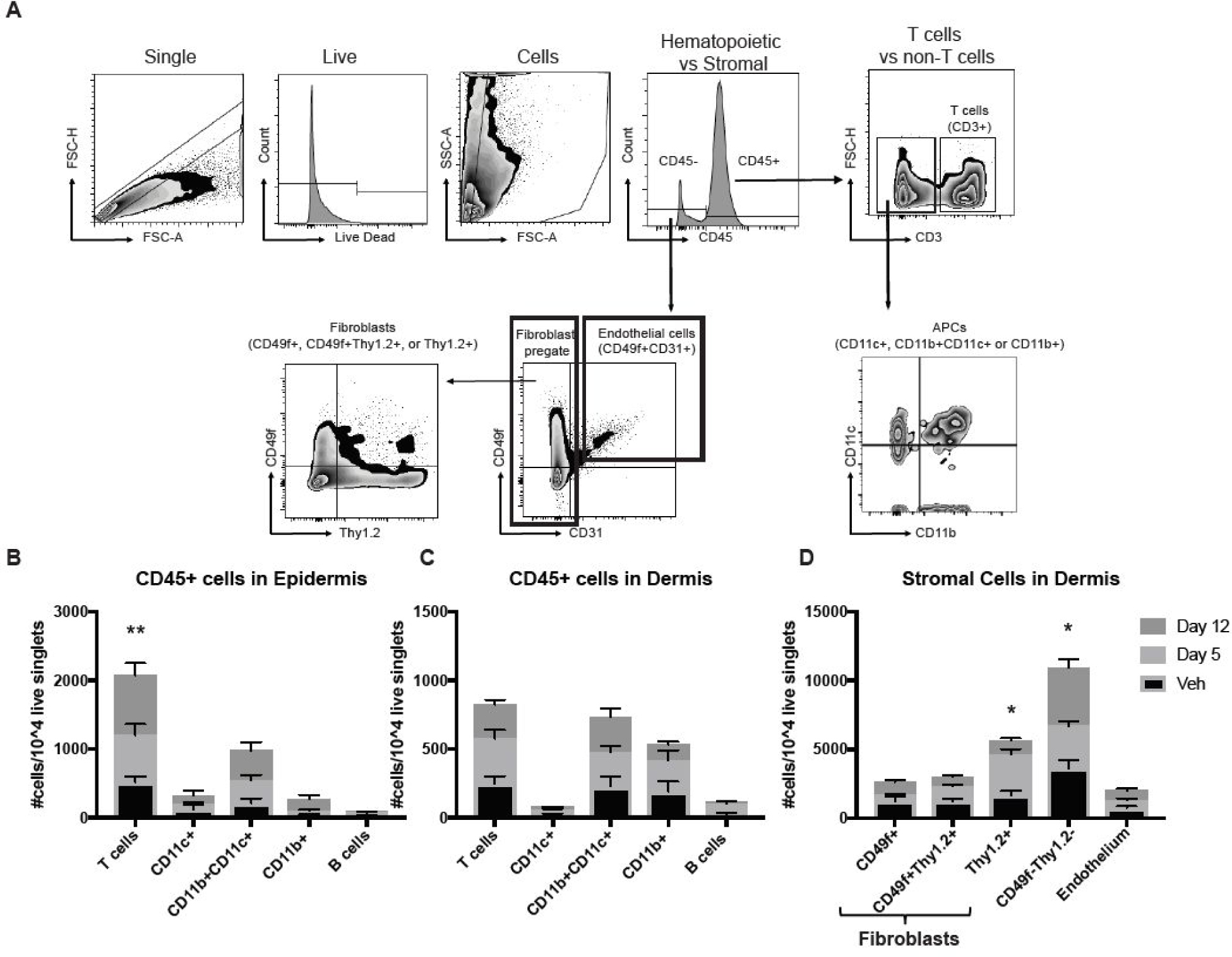
Characterization of inflammatory infiltrates and stromal cell populations in bleomycin-treated mice reveals increases in T cells and fibroblasts. (**A**) Sample gating strategy used to identify different cell types in the skin of bleomycin or vehicle treated mice. Normalized CD45+ inflammatory cell numbers per 10,000 live singlets were calculated for (**B**) epidermis and (**C**) dermis. (**D**) Normalized CD45-cell numbers per 10,000 live singlets were calculated and significant increases were detected in fibroblast populations in the dermis. (two-way ANOVA, epidermal T cells **p = 0.0079 vehicle vs. day 12, Thy1.2+ stroma *p = 0.0126 vehicle vs. day 5, CD49f-Thy1.2- stroma *p= 0.029 day 5 vs. day 12; n=14 vehicle, 10 day 5, and 14 day 12 mice pooled from 3-5 experiments).

We also assessed non-hematopoietic cells in the dermal compartment by pregating on CD45-cells. We used CD31+CD49f+ to identify endothelial cells as previously described (Gordon et al. 2010). We confirmed that Thy1.2 and CD49f surface markers identified fibroblast populations by performing cell sorting and qPCR for cell-specific transcripts, and found high levels of FSP1 and Col1a1 expression in Thy1.2+, CD49f+, and Thy1.2+CD49f+ populations (Fig S2). Interestingly, we found statistically significant increases in the number of Thy1.2+ (CD90+) fibroblasts (Fig 2D). We also found increases in the number of CD49f-Thy1.2- populations, which did not express FSP1 or Col1a1 by qPCR and likely represent other non-hematopoietic cells such as neurons, fat cells, etc. (Fig 2D). Taken together, these flow data help to characterize the inflammatory and skin-resident cells in the early subcutaneous bleomycin model.

### Specific cell populations significantly upregulate CXCL9 and CXCL10 in the skin following bleomycin injection

We and others have shown increased expression of CXCL9 and CXCL10 in lesional skin of morphea patients, particularly in inflammatory lesions and in the serum of patients with active disease (Mertens et al. 2018; O’Brien et al. 2017). To assess whether our mouse model recapitulated the gene signature we found in human patients, we used the REX3 mouse, which reports the expression of CXCL9 and CXCL10 with red or blue fluorescent proteins respectively (Groom et al. 2012; Richmond et al. 2017). Injection of PBS did result in basal chemokine expression, though this is to be expected after needle stick in the skin (Melikoglu et al. 2006). Treatment with bleomycin upregulated CXCL9 expression by 5 days, and statistically significantly increased the amount of dual CXCL9/10 production by 12 days (one way ANOVA p=0.0032 with Tukey’s post tests *p=0.0315, **p=0.0036, Fig 3A). CXCL9 and CXCL10 reporters were upregulated in the dermis and epidermis, with the highest upregulation in the dermis (Fig 3B & Fig S3). We found no upregulation of these chemokines in skin draining lymph nodes, indicating a skin-specific effect of the subcutaneous bleomycin injection (Fig S4). Of note, inflammatory skin conditions can result in leakage of chemokines into the blood, which is why we believe there is an increase in CXCL9 and CXCL10 in morphea patient serum. Therefore, we believe this model recapitulates the CXCR3 system as observed in human morphea.

**Fig. 3.**
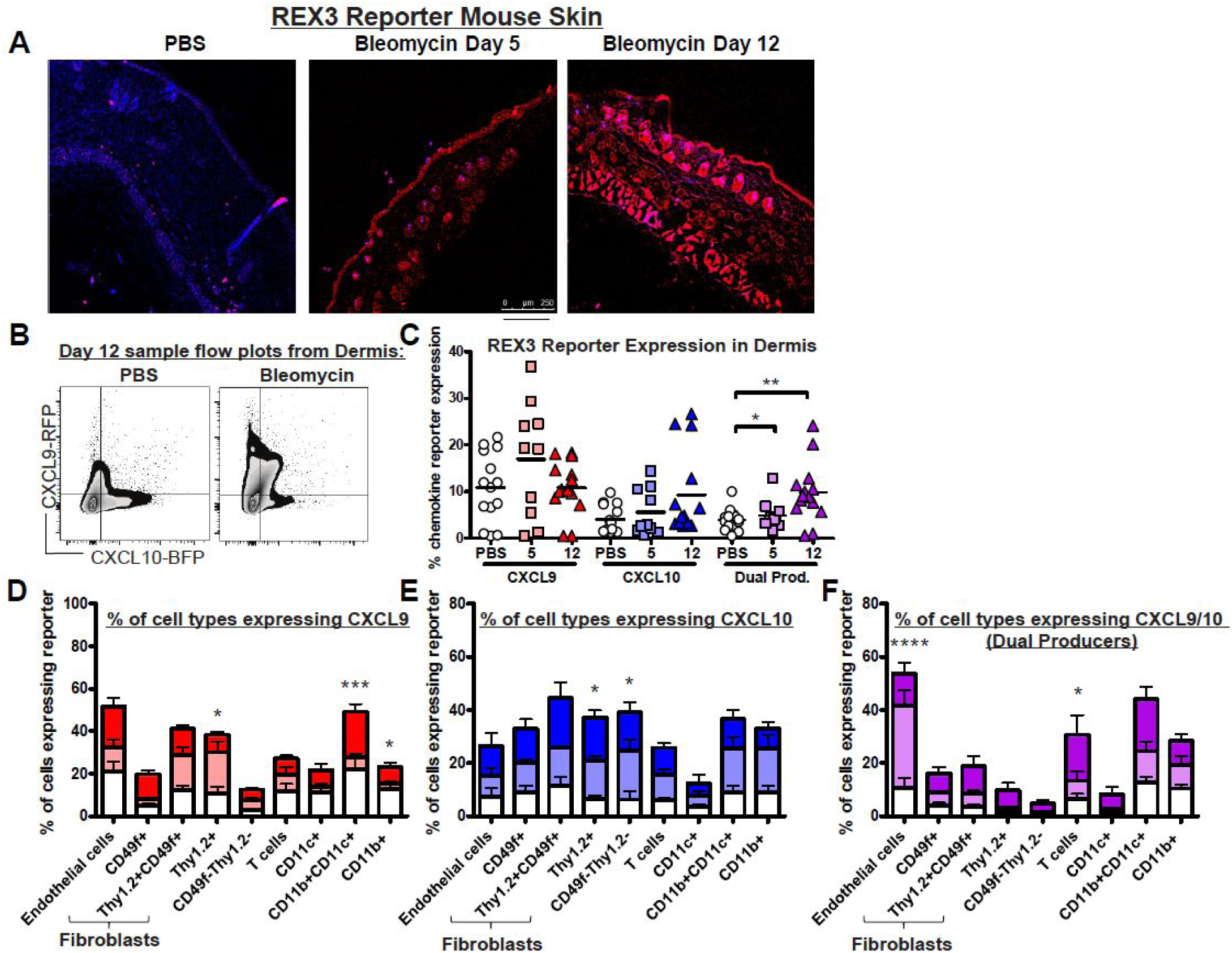
Chemokine reporter expression analysis in different cell types in the dermis following bleomycin administration reveals that fibroblasts, macrophages, and endothelial cells are key sources of CXCL9 and CXCL10. (**A**) Sample histology of skin from PBS or bleomycin treated mice (10x confocal microscopy). (**B**) Sample flow gating on CXCL9-RFP and CXCL10-BFP reporters in the dermis of vehicle or bleomycin-treated mice, pre-gated on live single cells. (**C**) Quantification of total chemokine produced at the indicated time points (one-way ANOVA of CXCL9 p=0.1667, CXCL10 p=0.0936, dual producers p=0.0032 with Tukey’s post tests: **p= 0.0036 vehicle vs. day 12, *p=0.0315 day 5 vs. day 12). To assess which cells were producing chemokines, dermis tissue was gated on live single cells, then cell types, followed by chemokine reporter to assess how many of each cell type was producing chemokines. (**D**) A significant number of CD11b+CD11c+ and CD11b+ macrophages and Thy1.2+ fibroblasts upregulated CXCL9 (two-way ANOVA p<0.0001 for cell type with Tukey’s post tests: CD11b+CD11c+ macrophages ***p=0.0005 vehicle vs. day 5 and **p=0.001 day 5 vs. day 12, CD11b+ macrophages *p=0.0406 vehicle vs. day 5, Thy1.2+ fibroblasts *p=0.0224 day 5 vs. day 12), whereas (**E**) Thy1.2+ and CD49f-Thy1.2- fibroblasts upregulated CXCL10 (two-way ANOVA p=0.0042 for cell type with Tukey’s post tests: Thy1.2+ fibroblasts *p=0.0469 vehicle vs. day 12, CD49f-Thy1.2- fibroblasts *p=0.0222 vehicle vs. day 5). (**F**) Endothelial cells and T cells upregulated both CXCL9 and CXCL10 (two-way ANOVA P<0.0001 for cell type with Tukey’s post tests: endothelial cells ****p<0.0001 for vehicle vs. day 5 and day 5 vs. day 12, T cells *p=0.0273 vehicle vs. day 12 and *0.0398 day 5 vs. day 12; n=14 vehicle, 10 day 5, and 14 day 12 mice pooled from 3-5 experiments).

We examined which cells in the skin upregulated CXCL9 and CXCL10 in REX3 mice following bleomycin treatment, and found that the ligands were upregulated by many cell types, though only specific subsets gained statistical significance (Fig 3D-F, Fig S3 & Fig S4). Fibroblasts contributed the most to CXCL9 and CXCL10 production, similar to our previous finding of keratinocyte contribution of these ligands in the epidermis during vitiligo (Richmond et al JID 2017). A significant number of APCs, specifically CD11b+CD11c+ and CD11b+ macrophages, upregulated CXCL9 production over time. A significant number of Thy1.2+ fibroblasts upregulated CXCL9 and CXCL10, and CD49f-Thy1.2- cells upregulated CXCL10 (Fig 3D & E). A significant number of endothelial cells and T cells upregulated both CXCL9 and CXCL10 (Fig 3F). In the epidermis after bleomycin treatment, overall chemokine expression was not significantly increased, though keratinocytes (CD45-) and Langerhans cells (CD11b+CD11c+) did appear to upregulate the chemokines, likely in response to the needle stick. Taken together, these data track the production of chemokine expression in acute bleomycin administration in the skin, which mirrors similar findings in human morphea patient skin (O’Brien et al. 2017).

### Bleomycin induces CXCL9 expression in mouse dermal cultures

To determine whether CXCL9 and CXCL10 expression in this mouse model are induced directly by bleomycin or if their expression is dependent on inflammatory cytokine production downstream of bleomycin-induced DNA damage, we cultured REX3 mouse dermis monolayers with 1.8μg/mL bleomycin (Nicolay et al. 2016) or with 1ng/mL IFNγ + 100ng/mL TNF (Yamaguchi et al. 1991), and measured REX3 reporter expression with a plate reader. Bleomycin was able to directly induce CXCL9 production in the absence of detectable IFNγ secretion as assayed by ELISA of culture supernatants. In contrast, IFNγ + TNF were needed to induce CXCL10 production (Fig 4).

**Fig. 4.**
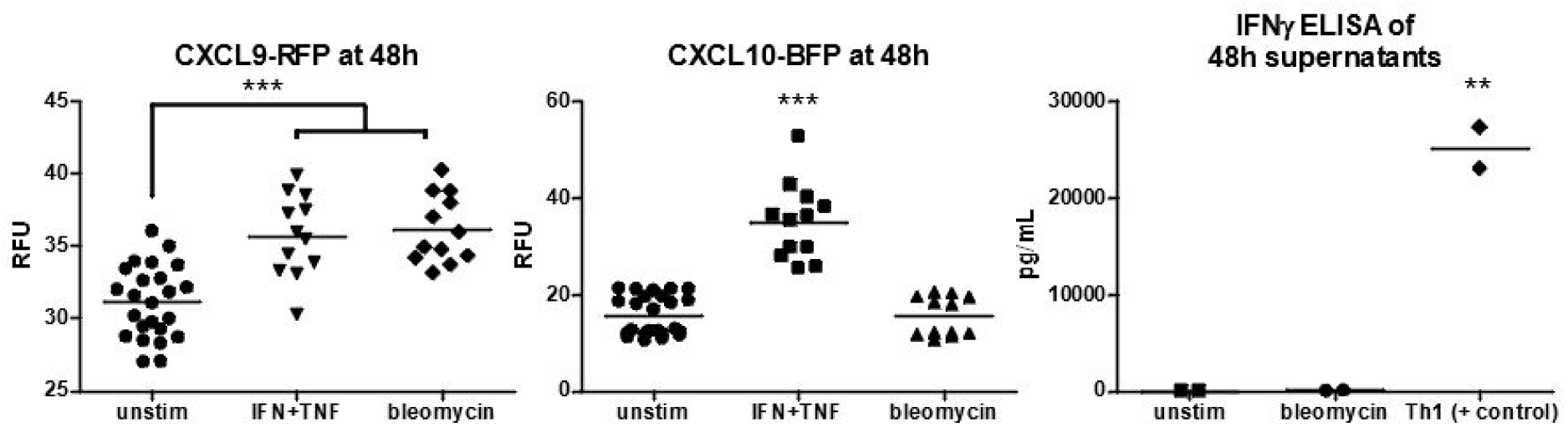
Bleomycin directly induces CXCL9 but not CXCL10 in dermis cultures. Dermis tissue was isolated from REX3 mice and cultured to confluency prior to 48h stimulations as indicated. (**A**) 1ng/mL IFNγ + 100ng/mL TNF (IFN+TNF) and 1.8μg/mL bleomycin significantly induce CXCL9 reporter induction compared to unstimulated controls (unstim). (**B**) Only IFN+TNF were able to induce CXCL10 production (n=2 experiments pooled, each dot represents one well/replicate, one-way ANOVAs with Tukey’s post tests p<0.0001). (**C**) Incubation of dermis monolayers with bleomycin did not induce detectable release of IFNγ (Th1 = T helper type 1 mouse T cells, positive control; one-way ANOVA with Tukey’s post test p= 0.0011).

### CXCL9 drives fibrosis in mouse morphea

We wanted to further examine the role of CXCR3 ligands in our morphea model to determine if they are also involved in pathogenesis, as mouse models of intravenous/intraperitoneal bleomycin to model systemic sclerosis have revealed a role for CXCL9 and CXCL10 in formation of early lung lesions (Tager et al. 1999). We tested CXCL9, CXCL10 and CXCR3-deficient mouse strains in the bleomycin morphea model. We found that CXCL9-deficient animals were protected from bleomycin-induced fibrosis as determined by dermal/epidermal thickness (*p=0.0119; Fig 5A). CXCR3-deficient animals also showed a similar degree of protection from fibrosis, although in this group the p value was 0.0568 compared to WT mice for a two-tailed test (p=0.0284 for one-tailed test of WT vs CXCR3KO). In contrast, CXCL10-deficient mice developed dermal thickening similar to control animals. Of note, CXCL10 has previously been shown to inhibit fibrosis in the lung in bleomycin-induced models of systemic sclerosis (Jiang et al. 2010; Tager et al. 2004), and therefore may actually play a protective role in the model. We confirmed the effect using hydroxyproline assay for collagen content.

**Fig. 5.**
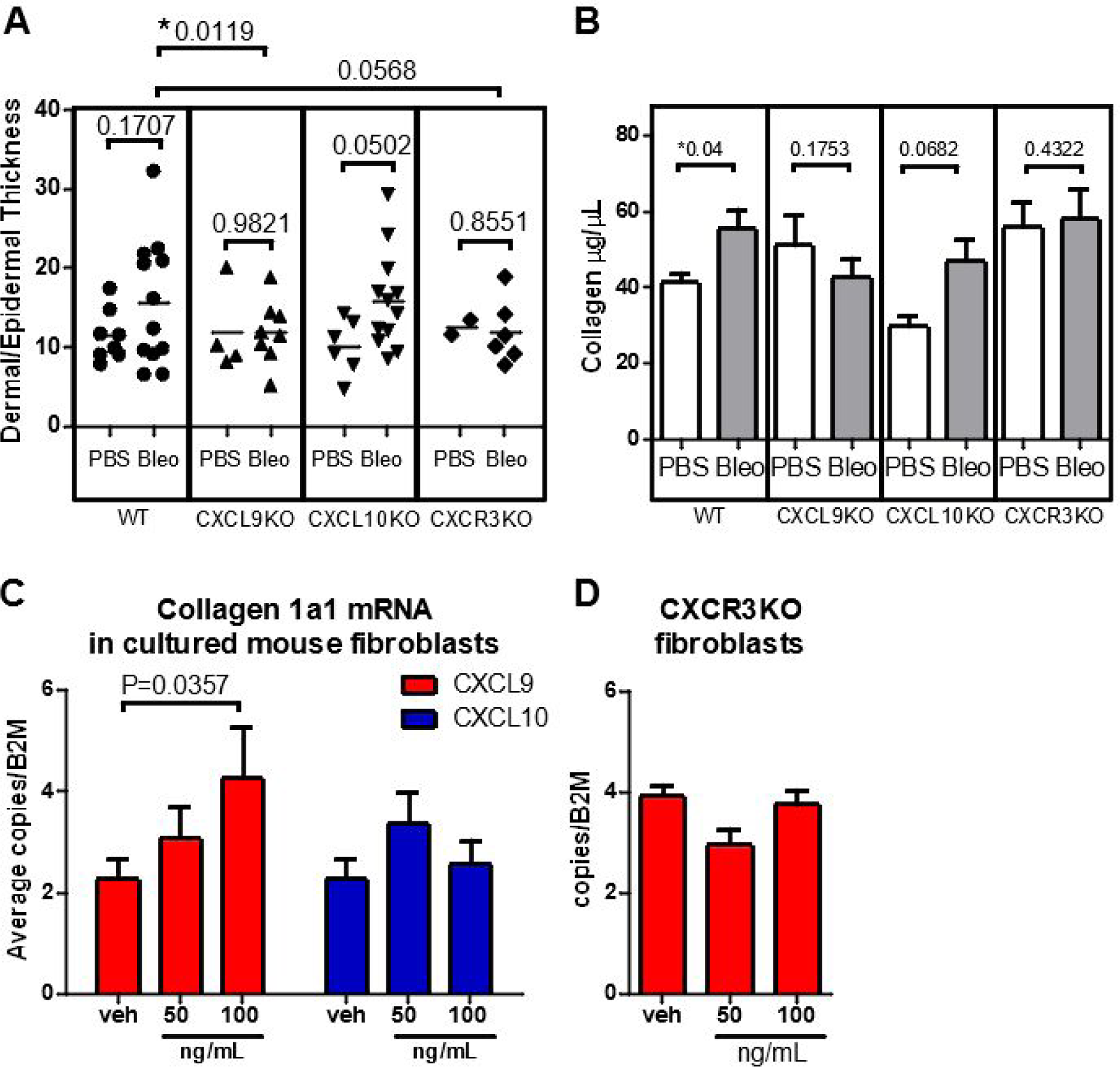
CXCL9-deficient animals are protected from bleomycin-induced fibrosis, and *in vitro* treatment of mouse fibroblasts with CXCL9 induces collagen 1a1 transcription. (**A**) Dermal:epidermal thickness ratios from different genotypes of mice treated with bleomycin reveals CXCL9KO and CXCR3KO mice had less thickening compared to WT controls (each dot represents a biopsy site; one-way ANOVA p=0.0295, posttests ns; t test of WT vs. CXCL9KO p=0.0119, WT vs. CXCR3KO p=0.0568; other t tests of PBS vs Bleo for each genotype are indicated). (**B**) Hydroxyproline assays for collagen content revealed significant increases in fold of collagen in the skin of WT mice treated with bleomycin and a trend toward significant increases in CXCL10KO mice treated with bleomycin, whereas CXCL9KO and CXCR3KO mice exhibited no significant differences when compared to vehicle controls (t tests for PBS vs Bleo treatment of each strain significant as indicated; t test comparisons of basal collagen by strain as follows: WT PBS vs. CXCL9KO PBS p=0.2642, WT PBS vs. CXCL10KO PBS p=0.0153, WT PBS vs. CXCR3KO PBS p=0.0565; n=5 WT PBS, 11 WT Bleo, 5 CXCL9KO PBS, 10 CXCL9KO Bleo, 3 CXCL10KO PBS, 11 CXCL10KO Bleo, 5 CXCR3KO PBS, and 11 CXCR3KO Bleo mice pooled from 2-3 separate experiments). (**C**) CD45-CD31-fibroblasts were sorted from naïve WT mouse dermis and were cultured for 5-7 days prior to treatment with recombinant CXCL9 or CXCL10, and were then assessed for collagen 1a1 mRNA production. CXCL9 dose-dependently increased Col1a1 mRNA in cultured fibroblasts whereas CXCL10 did not (5 pooled experiments, assayed in duplicate or triplicate, with technical duplicates for qPCR; t test vehicle vs 100ng/mL CXCL9 *p=0.0357). (**D**) CXCR3KO mouse fibroblasts were sorted and cultured for 5-7 days prior to assay as in panel C. CXCR3KO fibroblasts did not increase Col1a1 mRNA in response to CXCL9 (2-3 pooled experiments, assayed in duplicate or triplicate, with technical duplicates for qPCR).

The mice did exhibit basal differences in collagen content based on strain (Fig 5B) and sex (Fig S5 and as previously described (Arai et al PLOS One 2017)). Nevertheless, CXCL9KO and CXCR3KO mice did not exhibit increased collagen content compared to PBS injected controls, whereas WT animals exhibited significantly increased collagen content and CXCL10KO mice trended towards increased collagen following bleomycin treatment (Fig 5B). These data indicate that CXCL9 drives bleomycin-induced skin fibrosis and a morphea-like phenotype in mice, whereas CXCL10 is dispensable and may even play an anti-fibrotic role in light of the lack of significant protection in CXCR3-deficient animals.

### CXCL9 directly induces transcription of collagen 1a1 in mouse fibroblasts

To further investigate the different effects of CXCL9 and CXCL10 in fibrosis, we isolated live single CD45-CD31-fibroblasts from healthy mouse skin via cell sorting and cultured them in vitro for 5-7 days prior to treatment with recombinant CXCL9 or CXCL10. CXCL9 treatment induced dose-dependent increases in collagen 1a1 mRNA whereas CXCL10 did not (Fig 5C). CXCR3 deficient fibroblasts did not exhibit increased collagen production in response to treatment with CXCL9 (Fig 5D).

## Discussion

We have previously reported that CXCL9 serves as a biomarker of disease activity in morphea (O’Brien et al. 2017). However, no studies have examined immune dysregulation in morphea pathogenesis, and there is no validated animal model of morphea. To address this gap, we validated a murine model that replicates the features of early inflammatory morphea, and examined the role of the CXCR3 ligands in the development of morphea-like skin lesions in this model. We found increases in inflammatory cell populations as well as fibrosis, and histology from mice mirrors that seen in human skin. Taken together, our results support the use of the early subcutaneous bleomycin mouse model for preclinical studies in morphea and underscore the important role of CXCL9 in the development of skin fibrosis.

Using the REX3 reporter mice, we found that CXCL9 and CXCL10 are upregulated by a number of different populations within the skin following bleomycin treatment. Similar to our previous observations of keratinocytes and immune cells in the epidermis during vitiligo, we found that stromal cell populations contribute the bulk of chemokine production whereas antigen presenting cells have a much higher MFI of chemokine and therefore contribute the most on a per-cell basis (Richmond et al. 2017). Previous studies have shown that fibroblasts express Thy1 and CD49f (Korosec et al. 2019), and we found enrichment of fibroblast-specific transcripts in these cell populations. However, CD49f can also be expressed on stem-like cells in the dermis, so further characterization of fibroblast populations in the bleomycin mouse model are warranted. It will also be important to determine which populations are responsible for the fibrotic phenotype, and what their human homologues are. Additionally, we saw an increase in the number of CD45-CD49f-Thy1.2- cells, and characterization of these populations in mouse models of fibrosis

We found that fibrosis in our mouse model of morphea is dependent on CXCL9 but not CXCL10. This contrasts with other autoimmune/autoinflammatory conditions that have been shown to be dependent on CXCL10 but not CXCL9, including vitiligo (Rashighi et al. 2014), experimental autoimmune encephalomyelitis (EAE, (Fife et al. 2001)), and diabetes (Schulthess et al. 2009). The importance of CXCL9 as the sole ligand for driving fibrosis is in agreement with studies of renal fibrosis (Menke et al. 2008), but contrasts with a study of liver fibrosis showing that CXCL9 protects against fibrosis (Wasmuth et al. 2009). The conclusions in the liver fibrosis study were based upon a CXCR3-deficient mouse model and thus do not rule out antifibrotic contributions of CXCL10. In the context of pulmonary fibrosis, CXCL10 has been shown to play an anti-fibrotic role: specifically, CXCL10 binding to syndecan-4 inhibits migration of lung fibroblasts (Jiang et al. 2010; Tager et al. 2004). Gamma delta T cells have been identified as a key source of CXCL10 for attenuation of lung fibrosis (Pociask et al. 2011), and though gamma delta T cells are present in mouse skin (Itohara et al. 1993), they are notably absent in human skin (Vroom et al. 1991).

Differential signaling effects of CXCL9, CXCL10 and CXCL11 have been investigated in the context of MAPK, PI3K and calcium flux, and it is known that they bind different regions of CXCR3 (Colvin et al. 2004). Another group studied the effects of CXCL9, CXCL10 and CXCL11 on intestinal myofibroblasts and found differential signaling effects, though they found no detectable surface CXCR3 indicating either a variant or a different receptor for CXCR3 ligands in these cells (Kouroumalis et al. 2005). Studies of skin fibroblasts in models of wound healing and dermal differentiation have found that fibroblasts express CXCR3 (Kroeze et al. 2012; Yates et al. 2010; Yates et al. 2009; Yates et al. 2007). Nevertheless, there could be differences in fibroblast responses to CXCR3 ligands based upon their tissue of origin. Thus, the dichotomy of CXCL9 and CXCL10 effects in our model is likely due to skin fibroblast-intrinsic effects.

Limitations of our study include the need for a deeper understanding of the CXCL9-dependence of the disease, including signaling pathway activation in fibroblasts that permits collagen 1a1 production. Future studies are planned to focus on the signaling cascades involved in CXCL9 versus CXCL10 responses in fibroblasts. Taken together, these studies indicate that acute intradermal bleomycin injection may be a model for inflammatory and fibrotic morphea, and that this disease process is dependent on CXCL9 and its receptor CXCR3. These may be attractive targets for future morphea treatment options.

## Materials and Methods

### Study design

The objectives of this study were to characterize acute subcutaneous bleomycin injection as a model for inflammatory morphea, and to determine whether CXCL9, CXCL10 and CXCR3 are involved mechanistically in morphea in addition to their previously defined roles as biomarkers of disease. Our hypothesis was that the CXCR3 chemokine system is responsible for T cell infiltration into skin and was required for subsequent fibrosis. This hypothesis was formed on the basis of previous studies we have conducted in different animal models. We were surprised to find that CXCL9, and not CXCL10, was primarily driving disease in our model. Original sample sizes for mouse studies were based on our prior sample size determination for vitiligo model studies (see for example Richmond et al STM 2018) with a power of 0.8 and a significance level of 0.05, though we found that these numbers appeared to be under-powered in the morphea model, which required additional animals. Of note, we used animals conservatively from our colony, and report the numbers we were able to achieve without additional breeding. Replicate experiments were performed two to five times, as indicated in the figure legends.

Mice used for these studies were on the C57BL/6J (B6) background. Both male and female mice were assessed in accordance with the NIH policy on including both sexes in research studies. We did find that, in accordance with previous reports, female mice and fibroblasts appeared to have an enhanced response to bleomycin-induced collagen production, though we report aggregate data here. Fibrosis was assessed using a ratio of epidermal:dermal thickness on Trichrome stained skin sections to avoid error if tissue sections were not cut precisely perpendicularly (i.e. rather than just measuring dermal thickness). Hydroxyproline assays were conducted by our collaborators in separate laboratory spaces in a blinded fashion to corroborate our data. For assessment of inflammatory infiltration, mouse epidermis, dermis and lymph node samples were run on the flow cytometer on a high flow rate with a time cutoff of 60s per sample, and all cell numbers were normalized to live single cells as previously described.

### Human skin samples

Morphea lesional skin histology was obtained from a dermatopathologist’s biorepository (A.D.) from a skin biopsy from a 4 year old female diagnosed with morphea. Healthy skin histology was obtained from discarded skin tissue from a panniculectomy surgery under an institutional review board (IRB) approved protocol at UMMS (J.E.H.).

### Mice

All mice were housed in pathogen-free facilities at UMMS, and procedures were approved by the UMMS Institutional Animal Care and Use Committee and in accordance with the National Institutes of Health (NIH) Guide for the Care and Use of Laboratory Animals. Mice used for these studies were on the C57BL/6J (B6) background or a mixed 129 × C57BL/6 background that had been backcrossed to B6 for more than 10 generations. Age and sex-matched mice were used, and both male and female mice of all strains were tested to avoid gender bias. Replicate experiments were performed two to five times.

The following strains were used in the morphea model: C57BL6/J (The Jackson Laboratory stock no. 000664), Cxcl9-/- (B6.129S4-Cxcl9tm1Jmf, provided by J. M. Farber, National Institute of Allergy and Infectious Diseases, Bethesda, MD), Cxcl10-/- (B6.129S4-Cxcl10tm1Adl/J, The Jackson Laboratory, stock no. 006087), Cxcr3-/- (B6.129P2-Cxcr3tm1Dgen/J, The Jackson Laboratory, stock no. 005796) and REX3 (provided by A.D. Luster, Massachusetts General Hospital/Harvard Medical School).

### Bleomycin-induced mouse model of acute morphea

Bleomycin (Sigma-Aldrich) solution was prepared by dissolving bleomycin in PBS to a final concentration of 1mg/mL. Two locations on the dorsal side of the mouse were shaved for subcutaneous injections. Mice received injections of 50 µL of bleomycin treatment solution or PBS vehicle at each site (total 100μg/mouse) for 12 days. At the end of the experiment, all knockout and WT control mice were sacrificed, and half of each site was frozen for collagen and RNA analysis, and the other half was prepared for histological analysis. For analysis of REX3 mice, flow cytometry was performed as a readout to determine which cells were producing CXCL9 and CXCL10.

### Histological Analysis of Fibrosis

The skin from the injection sites was fixed using 4% paraformaldehyde prior to paraffin embedding (UMMS DERC core facility). Samples were sectioned and stained with trichrome to visualize collagen deposition or H&E to visualize inflammatory infiltrates. Images were taken with an Olympus BX51 microscope at 10x, and epidermal and dermal thicknesses were measured with Nikon NIS Elements BR software version 3.10. The ratio of the dermal to epidermal thickness was calculated for each skin sample to control for variances in skin sampling (i.e. cutting on a bias instead of perpendicularly).

### Flow cytometry

Tissues were harvested at the indicated times. Lymph nodes were mechanically disrupted, and back skin was incubated with 50U/mL (approximately 50mg/mL) Dispase II (Roche) for 1h at 37°C. Epidermis was removed and mechanically dissociated using 70μm filters and syringe plungers. Dermis was incubated with 1mg/mL collagenase IV and 2mg/mL DNAse I (Sigma Aldrich) for 1h at 37°C before mechanical dissociation. All samples were filtered prior to staining. Naïve REX3 mice were taken down at each time point and used to set gates for chemokine reporter analysis, and UltraComp beads (eBioscience) stained with Alexa Fluor 555 antibodies (Invitrogen) or Pacific Blue antibodies (Biolegend) were used to set up compensation for RFP and BFP. All murine flow cytometry samples were blocked with 2.4G2 and stained with LiveDead Blue (Invitrogen, 1:1000). Please see table S1 for lists of antibodies and clones used for these studies.

### Dermis and Fibroblast *in vitro* treatments

Dermis tissue was isolated from different skin sites from eight week or older age and sex matched animals as previously described (Richmond et al. 2017). Briefly, epidermis was removed by incubating skin with 5 (tail) or 50 (ear, back) mg/mL Dispase II enzyme (Roche) at 37°C for 1h or overnight at 4°C. Pooled dermis tissue was digested for 45min-1h at 37°C using 1mg/mL collagenase IV and 2mg/mL DNAse I (Sigma Aldrich), and tissue was mechanically dissociated using the blunt end of a syringe. Samples were filtered through 70μM mesh and were centrifuged at 330g for 10min in FACS buffer (1% FBS in PBS) containing 1mg/mL DNAse I.

For REX3 dermis cultures, the cell pellets were resuspended in complete DMEM (10% FBS, 1% Pen/strep, Corning) and cultured overnight in tissue culture dishes. The next day, debris and old media was aspirated off and fresh complete DMEM was added to the adherent cell culture. Cells were grown to a monolayer (3-4 days), then were trypsinized and plated in 96 well flat bottom plates for 2 days to re-form confluent monolayers prior to use in stimulation assays. For chemokine reporter assays, REX3 dermis monolayers were cultured with 1ng/mL IFNγ + 100ng/mL TNF (R&D Systems) or 1.8μg/mL bleomycin for 48h. Plates were read with a Molecular Devices Spectramax Gemini XPS plate reader with Softmax Pro version 5 software using the following ex/em filters and cutoffs: 405/460nm with 475nm cutoff, 544/555nm with 590nm cutoff.

For cell sorting of fibroblasts, dermis pellets were stained with anti-CD45 APC and anti-CD31 PE-Cy7 and all double negative singlets were collected by cell sorting with a FACS Aria. These cells, essentially all 4 phenotypes of mouse skin fibroblasts, were cultured in DMEM containing 10% FBS and 1X pen/strep for 5-7 days with one split. Cells were grown to 80% confluency and were used in collagen production assays. For collagen mRNA assays, 50-100ng/mL recombinant mouse CXCL9 or CXCL10 (R&D Systems) or vehicle control (PBS) were added to WT fibroblasts for 3-4h, and the cells were harvested by trypsinization and centrifugation. Pellets were snap frozen and were assessed for protein or RNA analysis as described below.

### Hydroxyproline assay

To quantify the amount of collagen in mouse skin specimens, hydroxyproline content was measured as previously described (Santos et al. 2009). Briefly, Dorsal skin tissues were digested overnight at 110°C in 1 ml of 6 N HCl. Samples (10 μl) were transferred to 96-well plates and mixed with 100 μl chloramine T solution (1.4% chloramine T, 10% isopropanol, 0.5 M sodium acetate, pH 6.0) for 15 minutes at room temperature, followed by 100 μl Erlich’s solution (14.9% p-dimethylaminobenzaldehyde, 70% isopropanol, 20% perchloric acid) and incubated at 65°C for 15 minutes. Absorbance was measured at 540 nm. Collagen content was calculated by comparison to a standard curve generated with cis-4 hydroxy-l-proline (0.01–110 μg/ml; Sigma-Aldrich), using the conversion factor of 1 μg hydroxyproline, corresponding to 6.94 μg collagen.

### qPCR

RNA was isolated from mouse back skin or fibroblast cell pellets using RNeasy kits (Qiagen). cDNA was prepared using iScript kits (Biorad), and qPCR was performed with SYBR green kits (Biorad). Mouse primers used were: Col1a1-F GAGCGGAGAGTACTGGATCG, Col1a1-R GCTTCTTTTCCTTGGGGTTC (Ito et al. 2010); B-actin-F GGCTGTATTCCCCTCCATCG and B-actin-R CCAGTTGGTAACAATGCCATGT (Rashighi et al 2014); and B2M-F CCGAACATACTGAACTGCTACGTAA and B2M-R CCCGTTCTTCAGCATTTGGA (Groom et al. 2012).

### Statistics

All statistical analyses were performed with GraphPad Prism software. Dual comparisons were made with unpaired Student’s t test. Groups of three or more were analyzed by ANOVA with Tukey’s (to compare all groups) or Dunnett’s (to compare to control group) posttests as indicated. For REX3 reporter studies, two-way ANOVAs with simple means effects were used to compare different cell types at different time points with Tukey’s post tests. P values < 0.05 were considered significant.

## Data Availability

Primary data are available upon request. Materials (REX3 mice and CXCL9-deficient mice) were obtained through MTAs with Massachusetts General Hospital and the National Institutes of Health.

## Supplementary Materials List

Fig. S1. Sample lung histology from mice treated with subcutaneous bleomycin reveals the absence of lung fibrosis.

Fig. S2. Enrichment of fibroblast-specific transcripts in sorted dermal populations.

Fig. S3. Chemokine expression in the skin draining lymph nodes of REX3 mice treated with subcutaneous bleomycin.

Fig. S4. Chemokine expression in the epidermis of REX3 mice treated with subcutaneous bleomycin.

Fig. S5. Sex differences in baseline collagen production in hydroxyproline assay.

Table S1. Flow antibodies and clones used for this study.

## Acknowledgments

We thank M. Ahmed Refat, C. Hsueh, C. Torok, E. Mirizio, W. Damsky and B. King for insightful comments on the manuscript, A. Luster for REX3 mice and J. Farber for CXCL9-deficient mice.

## Funding

Supported by a Calder Research Scholar Award from the American Skin Association, a Career Development Award from the Dermatology Foundation (to J.M.R.), NIH awards AR09114 and AR07302 (to J.E.H.). Flow cytometry and confocal microscopy equipment used for this study is maintained by the UMMS Flow Cytometry Core Facility and Morphology Core Facility, and tissue sectioning and pathology services are maintained by the UMMS DERC Morphology Core.

## Author contributions

J.M.R., D.P., T.W., M.G., C.J.G., & K.D. performed experiments and analyzed data. A.D. provided patient samples and management. H.J., J.E.H. & J.M.R conceived the study, designed experiments and provided study oversight. D.P. & J.M.R. wrote the manuscript, and all authors approved the final version.

## Competing interests

JMR and JEH are inventors on patent application #62489191, “Diagnosis and Treatment of Vitiligo” which covers targeting IL-15 and Trm for the treatment of vitiligo. JEH is scientific founder of Villaris Therapeutics, Inc, which is focused on developing treatments for vitiligo. JMR and JEH are inventors on patent application #15/851,651, “Anti-human CXCR3 antibodies for the Treatment of Vitiligo” which covers targeting CXCR3 for the treatment of vitiligo.

## Data and materials availability

REX3 mice and CXCL9-deficient mice were obtained through MTAs with Massachusetts General Hospital and the National Institutes of Health, respectively.

## Notes

### Author Declarations

All relevant ethical guidelines have been followed and any necessary IRB and/or ethics committee approvals have been obtained.

Any clinical trials involved have been registered with an ICMJE-approved registry such as ClinicalTrials.gov and the trial ID is included in the manuscript.

